# Patient-reported respiratory outcome measures in the recovery of adults hospitalised with COVID-19: A systematic review and meta-analysis

**DOI:** 10.1101/2022.03.16.22272509

**Authors:** Sophie Middleton, Christos Chalitsios, Tricia. M. McKeever, Alex R. Jenkins, Charlotte. E. Bolton

## Abstract

**Background:** Acute COVID-19 clinical symptoms have been clearly documented, but long-term functional and symptomatic recovery from COVID -19 is less well described.

**Methods:** A systematic review and meta-analysis were conducted to describe patient-reported outcome measures (PROMs) in adults at least 8 weeks post hospital discharge for COVID-19. Comprehensive database searches in accordance with the PRISMA statement were carried out up till 31/05/2021. Data were narratively synthesized, and a series of meta-analyses were performed using the random-effects inverse variance method.

**Results:** From 49 studies, across 14 countries with between 2-12 months follow up, the most common persisting symptom reported was fatigue with meta-analysis finding 36.6% (95 % CI 27.6 to 46.6, n=14) reporting it at 2-4 months, decreasing slightly to 32.5% still reporting it at >4 months (95% CI 22.6 to 44.2, n=15). This was followed by dyspnoea. Modified MRC score (mMRC) ≥1 was reported in 48% (95% CI 30 to 37, n=5) at 2-4months reducing to 32% (95% CI 22 to 43, n=7) at 4 months. Quality of life (QOL) as assessed by the EQ-5D-5L VAS remained reduced at >4 months (73.6 95% CI 68.1 to 79.1, n=6). Hospitalisation with COVID-19 also resulted in persisting sick leave, change in scope of work, and continued use of primary and secondary healthcare.

**Conclusion:** The symptomatic and functional impact of COVID-19 continues to be felt by patients months after discharge from hospital. This widespread morbidity points towards a multi-disciplinary approach to aid functional recovery.

## INTRODUCTION

Since emerging in December 2019, COVID-19 has impacted people, society and healthcare worldwide. As of 15^th^ March 2022 there has been over 450 million reported confirmed cases and 6 million deaths. [1] Although there is a significant acute mortality the majority of patients with COVID-19 infection will survive. [2] Attention then shifts to consideration of long-term morbidity and its effects on society, the economy and healthcare. Previous coronavirus outbreak survivors have shown continued fatigue, persistent shortness of breath and reduced quality of life up to 12 months post illness. [3] Hospitalised pneumonia patients experience fatigue, cough, and dyspnoea during recovery with adverse impacts on functioning and healthcare utilisation. [4] Studies of early recovery have shown similar ongoing morbidity [5, 6] with continuing abnormal respiratory function in COVID-19 survivors. [7]

Often there is a mismatch between patients and clinicians on when recovery has been reached. A negative COVID-PCR test, discharge from hospital and resolution of pneumonia on thoracic imaging can all occur with a person experiencing ongoing symptoms and reduced functioning. [8] Patient reported outcomes (PROMs) are measurement tools that are a useful way for patients to provide relevant information on their health, including quality of life, symptoms, and day-to-day functioning to help provide information on their recovery. [9] This systematic review and meta-analysis aimed to investigate the respiratory symptomatic and functional recovery of patients hospitalised with COVID-19 more than 8 weeks after hospitalisation using PROMs.

## METHODS

This systematic review was registered with PROSPERO (CRD42021242134) [10] and followed PRISMA guidelines for reporting systematic reviews. [11]

### Search strategy

The following databases were searched on 31/05/2021 for relevant studies: Embase, PubMed/MEDLINE, Cochrane COVID-19 Study register and CINAHL. The top 500 most relevant results on Google scholar were also searched as well as references within relevant articles and reviews. The search strategy included synonyms for COVID-19, respiratory outcomes and relevant terms related to quality of life and functioning and was not subject to language restrictions (Supplementary table 1).

### Eligibility and data extraction

Following collation of search results and removal of duplicate citations, title and abstract screening was undertaken by one reviewer to eliminate obviously irrelevant papers. Full papers were reviewed in duplicate for selection by four reviewers (SM, CC, TM, AJ). Papers were included if they contained participants over 18 years old, had a sample size of at least 30 patients who had been hospitalised in the acute phase, incorporated patients that had a clinical or PCR diagnosis of COVID-19 and had a minimum follow up time of 8 weeks post hospital discharge. A cut off sample size of 30 patients was used to avoid the inclusion of low powered studies and case reports. Studies were excluded if their primary outcome measures were focussed solely on non-respiratory or functional outcomes such as olfactory, gustatory, or neurological outcomes of COVID-19. Studies that included both hospitalised and non-hospitalised patients were included if they reported the hospitalised cohort separately. Studies that recorded follow-up from symptom onset were included if follow-up occurred at least 10 weeks following symptom onset as average time to admission to hospital is 5-6 days and average length of hospital stay is 5-15 days depending on age. [12]

Data extraction was undertaken using a proforma by four reviewers (SM, CC, TM, AR) and data agreed for each paper by two reviewers. Where disagreement remained, a third reviewer was included (CEB or TM). The proforma included patient selection, sample size, demographics, and study design. Main outcomes collected were patient reported symptoms, quality of life scores, breathlessness scales, functional testing, return to work and use of healthcare since discharge. The quality of the included studies was determined using the Newcastle-Ottawa score and Downs and Blacks checklist. [13, 14]

### Statistical analysis

Narrative synthesis of evidence was conducted for all included studies. Extracted results were analysed for adequate numbers of each outcome reported to perform a meta-analysis. Studies were split into separate groups where participant subgroups had been reported separately and it was not possible to combine the data. Meta-analysis using random effects models was performed to allow for apparent heterogeneity among studies given the different population characteristics. When looking at the chronology of follow up there were 2 peaks around 3 and 6 months. To capture this the studies were grouped considering the time after discharge (2-4 months and > 4 months) and meta-analyses were performed for each group. The generic inverse variance method was used for pooling. Pooled estimates were calculated using proportions with 95% CI for symptoms, modified medical research council dyspnoea score (mMRC) and mean difference with 95% CI for 6 minute walk test (6MWT), Short physical performance battery (SPPB), and EQ5D-5L. The Dersimonian-Laird method was used to estimate the between-study variance *T*^2^. The percentage of variability in the effect sizes not caused by sampling error was tested by using the Higgins’ *I*^2^ test, with estimates of 25%, 50% and 75% indicating low, moderate, and high heterogeneity respectively. If a study reported follow-up at multiple time-points with the same cohort, only the first follow-up timepoint was used in the meta-analysis to avoid bias. All meta-analyses were conducted in R v4.0.3 using the “meta” _and “metafor” packages and all statistical tests were two sided and used a significance level of p < 0.05.

## RESULTS

The search strategy identified 7331 papers of which 49 studies were included in the study (Figure 1). [15-62]

**Figure 1:**
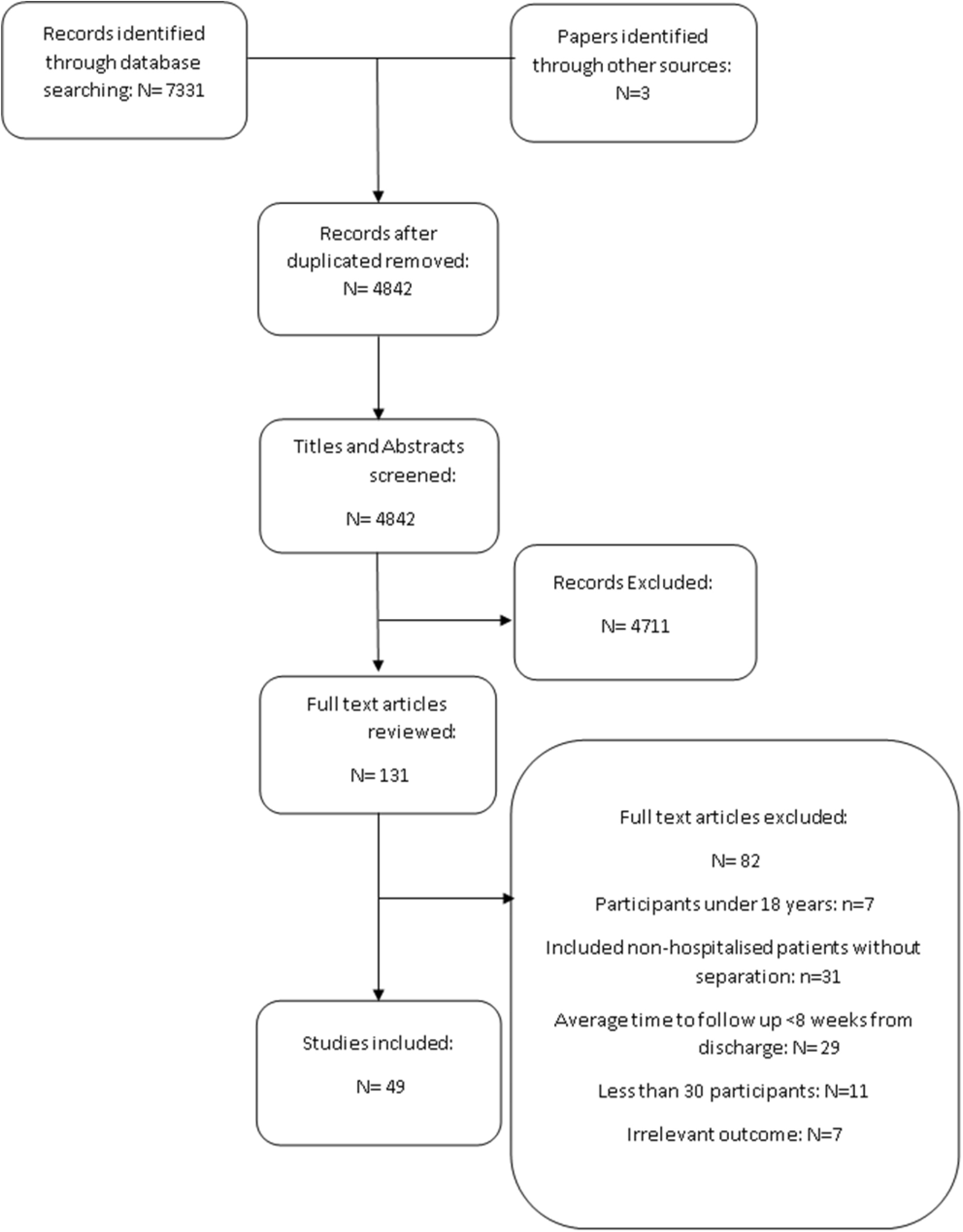
Paper selection process. Note some full text articles were excluded for multiple reasons.

### Study Characteristics

All studies were observational studies with follow-up time ranging from 2 to 12 months and originating from countries worldwide including Italy (11), China (10), UK (5), France (4), Spain (3), Switzerland (3), Canada (2), Netherlands (2), Norway (2), Belgium (2), USA (1), Denmark (1), Russia (1), Brazil (1), Japan (1). Participant numbers ranged from 36 to 2649, overall, 56% of participants were male. (Supplementary Table 2). Eight studies included only participants that were admitted to ITU (Intensive therapy unit). Outcome areas of interest ranged widely between the studies and the outcome tools were not standardised. See supplementary table 3 for the full list of individual outcome measures. Thirty-one studies reported patient respiratory/general symptoms, 33 respiratory or functioning PROMs, 27 health-related quality of life, 10 return to work and 6 healthcare utilisations. Most studies were of moderate quality scoring an average of 5.8(SD 1.6) out of a maximum of 10. The majority of studies were single site with no control group or adjustment for confounders (Supplementary Table 4). One patient cohort was reported at different timepoints over 2 publications. [33, 34]

### Patient-reported symptoms

Thirty-one papers reported on at least one respiratory or functional symptom and results from meta-analyses are reported in Table 1 (Forest plots Supplementary Figures 1 to 10). [15-17, 23, 24, 27-29, 32-34, 36-39, 41, 42, 44, 45, 47, 47-52, 57, 58, 61-63] The most prevalent reported symptom was fatigue (2 to 4 months; 36.6%; 95 % CI 27.6 to 46.6) which decreased slightly to 32.5% at >4 months (95% CI 22.6 to 44.2). This next most prevalent symptom at both times points was breathlessness.

**Table 1:** Meta-analysis of patient reported symptoms at 2-4 months and >4months after discharge.

| Symptoms | 2-4 months |  |  | >4 months |  |  |
| --- | --- | --- | --- | --- | --- | --- |
| | No of studies | Effect size (95% CI) | $I^2$ (%) | No of studies | Effect size (95% CI) | $I^2$ (%) |
| Breathlessness, % | 15 | 28.8 (21.5 to 37.5) | 94 | 11 | 22.0 (12.7 to 35.4) | 98 |
| Chest Pain , % | 6 | 5.1 (1.9 to 13) | 91 | 10 | 7.5 (4.9 to 11.3) | 95 |
| Cough, % | 14 | 13.3 (9.4 to 18.6) | 88 | 12 | 8.6 (5.4 to 13.7) | 95 |
| Fatigue, % | 14 | 36.6 (27.6 to 46.6) | 95 | 15 | 32.5 (22.6 to 44.2) | 98 |
| Myalgia, % | 4 | 11.8 (5.4 to 24.1) | 92 | 9 | 12.7 (6.4 to 24.4) | 99 |

### Respiratory and Functional outcomes

Thirty-three studies reported respiratory and functional outcomes across 18 different outcome measures. [15, 16, 20-25, 27, 28, 30-38, 40-43, 45, 47, 51-53, 55, 56, 59, 60, 63] The most common outcome measure reported was the mMRC breathlessness score (2 to 4 months; 48%; 95% CI 30 TO 67) reducing to 32% (95% CI 22 to 43) at >4 months (Table 2) (Forrest plots Online supplement Figure 11 to 21). The six-minute walk test(6MWT) was around 500 metres for both patients in 2 to 4 months and >4 months after discharge. [20, 30-35, 41, 45, 55, 60, 63]

**Table 2:** Meta-analysis of functional outcomes and health related quality of life at 2-4 months and >4months after discharge.

| Functional test | 2- 4 months |  |  | >4 months |  |  |
| --- | --- | --- | --- | --- | --- | --- |
| | No of studies | Effect size (95% CI) | $I^2$ (%) | No of studies | Effect size (95% CI) | $I^2$ (%) |
| **6MWT, m** | 6 | 501 (441.3 to 560.8) | 98 | 6 | 494.7 (457 to 532.4) | 98 |
| **SPPB** | - | - | - | 2 | 9.5 (8.4 to 10.6) | 92 |
| **mMRC 0, %** | 5 | 45 (31 to 60) | 89 | 7 | 67 (54 to 77) | 98 |
| **mMRC ≥1, %** | 5 | 48 (30 to 67) | 93 | 7 | 32 (22 to 43) | 98 |
| **EQ5D-Index** | 2 | 0.67 (0.60 to 0.74) | 72 | 5 | 0.74 (0.68 to 0.81) | 97 |
| **EQ5D-VAS** | 1 | - | - | 6 | 73.6 (68.1 to 79.1) | 98 |

Abbreviations: Six minute walk test (6MWT), Short physical performance battery (SPPB), modified medical research council (mMRC) dyspnoea score.

Three studies measured breathlessness according to the Borg scale at 9 weeks (51%) [20], 6 months (40%) [40] and 6 months (11%) [59] The full summary of outcomes not included in the meta-analysis can be seen in Supplementary Table 5 to 7.

### Health related Quality of life

Twenty-seven papers looked at patient quality of life using seven different outcome measures. The most common outcome measure used was the EQ-5D. [15, 18, 21, 27, 28, 32, 35, 38, 40, 42, 47, 51-54, 56, 57] The EQ-5D VAS score ranges from 0 to 100 with 100 being the best quality of life. The meta-analysis showed a EQ-5D-5L VAS score of 73.6(95% CI 68.1 to 79.1) at 4 months (Table 2).

Five studies [33, 34, 41, 43, 55] used the SF-36 score to assess quality of life at follow up. Similarly, marked out of 100, with 100 representing the best quality of life, participants reported being particularly limited in their role in society by their physical health problems (Range 23.7 to 68). [34, 55] (Supplementary Table 8). Gianella et al found Health-related quality of life to be below population average at 81 days follow up with an average score of 31±1.6 (50±10 expected average). [30] Using the 15-D HRQOL score, Gamberini et al found impairment in a participant’s breathing, usual activities, vitality, and sexual activity at 90 days. [26]

### Return to work

Ten studies reported on work resumption. [15, 19, 25, 27, 29, 33, 38, 40, 54, 62] Eight papers reported return to pre-illness job without work adjustments. [15, 19, 25, 27, 29, 33, 40, 62] This ranged from 20.5% back at work after 61 days [40] to 100% back at work after 3 months [62] At 61 days Monti et al [40] reported 48.7% of workers remaining on sick leave, whereas Latronico et al [33] reported 32% at 3 months and Evans et al [15] reported 17.6% at 5 months. Vaes et al found that on average participants missed 81.5% of work time at 3 month follow up decreasing to 60% at 6 months. [54] Of those at work 67.8% and 59.7% felt their work was impaired at 3 and 6 month follow up. [54] The proportion of participants who had a change in their scope of work or part time work ranged from 2.5% to 32%. [33, 38, 40, 62]

### Healthcare utilisation

Six studies reported on reattendance to healthcare providers following discharge. [25, 29, 38, 49, 50, 53] Rehospitalisation rate ranged from 8% to 28% at the time of follow-up across the 6 studies. Two studies found that 61.7% and 66.4% of participants had visited a GP between discharge and follow-up [38, 50] and 20.3% of participants had attended an emergency department by 3 months follow-up. [53]

## DISCUSSION

This comprehensive systematic review and meta-analysis investigating the respiratory and functional recovery of hospitalised adult patients with COVID-19 demonstrates over a third of participants continued to experience fatigue and 32% had an abnormal dyspnoea score over 4 months following hospital discharge. After 4 months the quality of life was below population average. [64]. Hospitalisation with COVID-19 resulted in persisting sick leave, change in scope of work and impairment at work up to 6 months post discharge and continued use of primary and secondary healthcare.

Other respiratory infections have shown similar ongoing respiratory and functional sequalae after discharge. [4, 65, 66] In a systematic review of community acquired pneumonia (CAP) at 6 weeks post infection, fatigue was seen in 42% (95% CI 10 to 74) and dyspnoea in 39% (95% CI 21 to 58). This was comparatively slightly higher than in our study at 36.6% (CI 27.6 to 46.6) and 28.8% (CI 21.5 to 37.5) respectively at 2-4 months follow-up. [4] In SARS, survivors continued to have reduced exercise capacity at 6 months post discharge with mean 6MWT 461 m (95% CI 450–473m) compared to 494.7m (95% CI 457 to 532.4) in our study at >4months. This was coupled with long-term respiratory, psychological sequalae and reduced health related quality of life. [65] Additionally, COVID-19 survivors had a higher burden and broader array of systemic sequelae than hospitalised seasonal influenza survivors at a mean follow up time of 150 days. This was coupled with an increased risk of death (HR= 1.51; 95% CI 1.30–1.76) and increased risk of needing outpatient care (HR= 1.12; 95% CI 1.08–1.17). [66]

The majority of patients testing positive for COVID-19 will not require hospital admission [67] and indeed persisting symptoms may occur in this group too with 47% of women and 33% of men having ≥1 symptom an average of 117 days post COVID-19 infection. [68] In our study between 8% to 28% of patients were re-hospitalised [25, 29, 38, 49, 50, 53] and 61.7% and 66.4% had visited their GP at the time of follow up [38, 50]. Up to 6 months after testing positive for COVID-19, 73% of non-hospitalised patients had visited their GP, a hospital outpatient clinic or been admitted to hospital. Healthcare usage was increased compared to matched COVID-19 negative patients. [69] Allocation of resources should be considered for management of COVID-19 recovery at all severity levels regardless of hospitalisation status.

We have reported the long-term respiratory and functional sequalae of COVID-19. Other studies have described a larger breadth of sequelae effecting mental health, neurological, gastrointestinal, metabolic, and cardiac systems. [66] WHO has recognised the urgent need for further understanding of the complexity of prolonged recovery from COVID, its wider economic and social implications and the need for new management pathways centred round multi-disciplinary care. [70] Care should involve rehabilitation, psychological intervention as well speciality management of physical health symptoms. [71] These follow-up clinics can also facilitate future research to improve our understanding and care of patient during recovery from COVID-19.

## Strengths and limitations

To the best of our knowledge this is the first meta-analysis looking at PROMs during prolonged recovery from COVID-19 hospitalised patients more than 8 weeks post discharge. It was conducted in accordance with the PRISMA guidelines. Studies varied widely in their study population selection and their outcome measures used. This limited the number of outcomes that could be used in the meta-analysis and contributed to the heterogeneity of results. The criteria for severe COVID-19 were defined differently between different countries/studies, this inhibited subgroup analysis based on this. There is a potential bias due to uncontrolled confounders and heterogeneity in observational studies. Due to insufficient data reporting in the original publications, we could not assess the effects of other factors, such as sex, age, and comorbidities in the proportion of symptoms. For future research, standardising PROMs used as well as classification of COVID-19 severity would aid translative research and collaborative work.

## CONCLUSION

We have shown that beyond 2 months since discharge respiratory symptoms, functional deficits and reduced quality of life are experienced by survivors of COVID-19 hospital admission. These results along with other emerging data, support the need for a multi-system, multi-disciplinary approach to support and aid functional recovery in these patients.

## Supporting information

Forest Plots

## Data Availability

All data produced in the present study are available upon request to the authors or in the supplementary materials.

## Authors contribution

S.M, C.E.B, and T.M.M designed the research and wrote the methodology. S.M performed the literature search. S.M, C.V.C, A.R.J and T.M.M were involved in the paper selection process. S.M, C.V.C, A.R.J and T.M.M extracted and checked data. C.V.C performed the meta-analyses. S.M drafted the paper. C.V.C, C.E.B, A.R.J and T.M.M critically reviewed and improved it.

## Acknowledgements

Thank you to Alison Ashmore, Senior Research Librarian at University of Nottingham libraries for help in devising a comprehensive literature search.

