## Supplementary tables for "Patient-reported respiratory outcome measures in the recovery of adults hospitalised with COVID-19: A systematic review and meta-analysis"

**Supplementary Material**

**Supplementary Table 1**. Database search strategy.

| Databases | Search terms |
| --- | --- |
| Embase,medline via Ovid and CINHAL, Cochcrane database individually | (Coronavirinae OR “Coronavirus infection” OR “severe acute respiratory syndrome” OR “SARS coronavirus” OR "Covid-19" OR "coronavirus disease 2019" OR “SARS-CoV-2”) AND (dyspn* OR "quality of life" OR cough* OR fatigue OR "return to work" OR "work resumption" OR "respiratory tract disease" OR ADL OR "exercise tolerance"/) AND (Recovery OR follow up OR treatment outcome OR outcome assessment OR patient-reported outcome OR " clinical outcome" OR prognosis) |
| Google Scholar (limited by 256 characters) | (dyspnoea OR "quality of life" OR cough OR “Work resumption” OR ADL OR “exercise tolerance”) AND (Recovery OR "follow up" OR "treatment outcome" OR "patient-reported outcome") AND ("Coronavirus" OR "Covid-19" OR "SARS-CoV-2") |

**Supplementary Table 2.** Study population inclusion and exclusion criteria and population characteristics. Key for time to follow up: * Mean, # Since symptom onset, $ Since diagnosis. Where subgroup populations were reported in one study and their demographics couldn’t be combined, those demographics are reported separately. NR= Not reported.

| Study | Year | Country | Study design | Median follow up from discharge (days) | Inclusion Criteria | Exclusion criteria | Participants | Age (years) | Male (%) | BMI | Length of stay (median days) | % requiring oxygen | % requiring ITU |
| --- | --- | --- | --- | --- | --- | --- | --- | --- | --- | --- | --- | --- | --- |
| Bellan et al | 2021 | Italy | Cohort | 98 | Over 18, consecutively admitted to hospital with laboratory or clinically confirmed covid-19 | Deceased patients | 238 | 61 (IQR 50-71) | 59.7 | NR | NR | 42.9 | 11.8 |
| Du et al | 2020 | China | cohort | 168 | laboratory-confirmed COVID-19 Patients consecutively admitted. | Patients who did not undergo a post-COVID-19 functional status scale (PCFS) interview at six-month follow-up after discharge or had a PCFS > 0 at baseline | 95 | 62 (IQR 53-69) | 52.6 | NR | NR | NR | NR |
| Fayol et al | 2021 | France | cohort | 168 | laboratory-confirmed COVID-19, requiring a hospital stay (>48 h) for a symptomatic COVID-19 pneumonia. | Pregnancy and inability to give consent or participation in an interventional study | 48 | 58 (SD 13) | 69.0 | 26 | NR | NR | 27 |
| Gonzalez et al | 2021 | Spain | cohort | 84 | Patients with positive SARS-CoV-2, older than 18 years, had ARDS, ICU admission | Hospital transfer, receiving palliative care, or severe mental disability that made it impossible to have pulmonary function tests after discharge | 62 | 60 (IQR 48-65) | 74.2 | 28.2 (IQR 25.4-32.6) | 14.5 (IQR 7-25.8) | NR | 100 |
| Huang et al | 2021 | China | cohort | 153 | laboratory confirmed COVID-19 who were discharged from hospital | Died, or unable to follow-due to psychotic disorder, dementia, or re-admission to hospital, unable to move freely, declined to participate, unable to be contacted, and living outside of Wuhan or in nursing or welfare homes | 1733 | 57 (IQR 47-65) | 52.0 | NR | 14 (IQR 10-19) | 68 | 4 |
| Latronico et al 3 months | 2021 | Italy | cohort | 84 | adult critically ill patients RT-PCR-confirmed SARS-CoV-2 infection and diagnosis of ARDS admitted to ICU | NR | 57 | 56 (SD 9.8) | NR | NR | 27.28 (SD 12.86) | NR | 100 |
| Latronico et al 6 months | 2020 | Italy | cohort | 168 | adult critically ill patients RT-PCR-confirmed SARS-CoV-2 infection and diagnosis of ARDS admitted to ICU | NR | 45 | 59 (IQR 54-64) | NR | NR | 27 (16-46) | NR | 100 |
| Lerum et al | 2020 | Norway | cohort | 83 | > 18 years, admitted for >8 hours with a discharge diagnosis of COVID-19 | living outside the hospitals’ catchment areas, inability to provide informed consent, or participation in the WHO trial Solidarity. | 103 | 59 (IQR 49-72) | 52.0 | 25.8 (IQR 23.8-29.6) | 6 (IQR 3-11) | 66 | 15 |
| Leth et al | 2021 | Denmark | cohort | 84 | Patients aged ≥18 years with a positive PCR test for SARS COV-2 who were hospitalized | transferred to another region | 49 | 58 (IQR 48-73) | 43.0 | 27.5 (*IQR* 24.5–32.7) | 6 (IQR 3-10) | 88 | 12 |
| Meng et al | 2020 | China | observational | 84, 168 | patients with COVID-19 pneumonia | NR | 46 | 45.29(SD 14.22), | 45.6 | 24.54 ± 3.67 | NR | NR | NR |
|  |  |  |  |  |  |  |  | 59.09 (SD 7.5) |  | 26.18 ± 2.76 |  |  |  |
|  |  |  |  |  |  |  |  | 61.73 (SD 15.15) |  | 24.94 ± 3.14 |  |  |  |
| Menges et al | 2021 | Switzerland | cohort | 196^$^ | aged >18 years, sufficient knowledge of the German language, and residing in the Canton of Zurich. | NR | 81 | 47 (IQR 33 - 58) | 50.0 | 24.8 | NR | NR | 10 |
| Miyazato et al | 2020 | Japan | cohort | 108* | Patients who were admitted due to COVID-19 | Patients who died during admission or their family were not invited to participate | 63 | 48.1 (SD 18.5) | 66.7 | 23.7 | 14 (SD 10.0) | 27.0 | NR |
| Monti et al | 2021 | Italy | cohort | 61 | all adult patients with COVID-19 ARDS admitted to ICU who received at least one day of invasive ventilation. | NR | 39 | 56 (SD 10.5) | 90.0 | 29 | 30 (IQR 23-44) | NR | 100 |
| Morin et al | 2021 | France | cohort | 113 | survival 4 months after hospital discharge, admitted to an ICU, > 18 years old, hospitalized > 24 hours because of COVID-19, and who had received a diagnosis of SARS-CoV-2 infection, or both. | death within 4 months after discharge, persistent hospitalization, end-stage cancer, dementia, nosocomial COVID-19 infection, and incidental positive SARS-CoV-2 RT-PCR result during a hospital stay | 478 | 60.9 (SD 16.1) | 57.9 | 28.8 | 9 (IQR 4-15) | NR | 29.70 |
| Munblit et al | 2021 | Russia | cohort | 217.5 | ≥18 years old, with both RT-PCR confirmed SARS-CoV-2 infection, and clinically confirmed infection, when the laboratory testing result is negative, inconclusive or unavailable. | NR | 2649 | 56 IQR (46-66) | 48.9 | NR | 14.6 (IQR 12-17.6) | 36.60 | NR |
| Qin et al | 2021 | China | cohort | 84 | NR | NR | 647 | 58 | 44.0 | 23.87 (sd 3.18) | 18 (SD 8) | NR | NR |
| Shah et al | 2021 | Canada | cohort | 81.9*^#^ | adults with COVID-19 hospitalised from March to May 2020 with laboratory-confirmed SARS-CoV-2 infection. | NR | 60 | 67 (IQR 54-74) | 68.0 | 25(23-29) | 10 (IQR 6-16) | 78 | NR |
| Sigfrid et al | 2021 | UK | cohort | 222 | Patients aged > 18 years, admitted to hospital with confirmed or highly suspected SARS-CoV-2 infection at 31 centres, who were discharged at least 90 days ago. | NR | 327 | 59.7 (IQR 51.7 - 67.7) | 58.7 | NR | 9 (IQR 5-20) | 79.20 | 39.8 |
| Smet et al | 2020 | Belgium | Cross-sectional | 74* | NR | NR | 220 | 53 (SD 13) | 62.0 | 28.1 | NR | NR | 18.6 |
| SuarezRobles et al | 2020 | Spain | cohort | 90 | consecutive non-probabilistic sampling, of patients discharged for COVID-19 from hospital, | Did not consent to participate | 134 | 58.53 (SD 18.53) | 46.3 | NR | NR | NR | 1.50 |
| Sykes et al | 2021 | UK | cohort | 113 | RT-PCR confirmed COVID-19 pneumonia | admission was unrelated to COVID-19 or mild symptoms and a normal chest x-ray. Those followed up under local frailty team | 134 | 58 (range 25-89) | 65.7 | 28.8 (range 17.3-49.8) | 7 (range 1-45) | 86.6% | 20.1 |
| Taboada et al | 2020 | Spain | cohort | 168 | all critically ill patients with COVID-19-induced ARDS admitted to the ICUs | NR | 91 | 65.5(SD 10.4) | 64.8 | NR | 35.21 (SD 21.09) | NR | 100 |
| van den Borst et al | 2020 | Netherlands | cohort | 71* | Patients who had been discharged after inpatient treatment for COVID-19 were consecutively invited to the aftercare facility. | NR | 97 | 57 (SD 10) | 68.0 | 27.2 (SD 3.2) | 8 (IQR 5-14) | NR | NR |
|  |  |  |  |  |  |  |  | 63 (SD 13) |  | 29.6 (SD 4.6) |  |  |  |
|  |  |  |  |  |  |  |  | 61 (SD 14) |  | 27.9 (SD 4.8) |  |  |  |
| Walle-Hansen et al | 2021 | Norway | cohort | 186 | patients aged >60 years, admitted hospital due to COVID19,still alive 180 days after hospital admission | Patients with SARS-CoV-2 without symptoms of COVID19 hospitalised for any other reasons | 106 | mean 74.3 (range 60–96 | 57.0 | NR | mean 11.6 (range 1-67) | NR | NR |
| Wong et al | 2020 | Canada | cohort | 91^#^ | Patients aged 18 years or older who were hospitalized laboratory-confirmed SARS-CoV-2 infection and able to complete surveys in English. | No exclusion criteria | 78 | 62 (SD 16) | 64.0 | NR | NR | NR | NR |
| Wu Qian et al | 2021 | China | cohort | 168 | patients who met the COVID-19 diagnostic criteria developed by the Ministry of Health of the People’s Republic of China, with diagnosis confirmed using nucleic acid testing. | Patients whose follow-up and physical examination data were missing, patients with chronic lung disease, or patients with mental illness who were unable to participate | 54 | 43.4 (SD 15.0) | 58.1 | NR | NR | NR | NR |
|  |  |  |  |  |  |  |  | 54.4(SD 13.6) | 60.9 |  |  |  |  |
| Xiong et al | 2020 | China | cohort | 168 | inpatients from 20 to 80 years of age diagnosed with COVID-19 according to WHO guidance and were discharged from the hospital | those with severe and complex underlying diseases or receiving invasive treatment, as well as women who were pregnant or breastfeeding | 538 | 52 (IQR 41-62) | 45.5 | NR | NR | NR | NR |
| Zhao et al | 2020 | China | cohort | 97 | All adult patients, diagnosed with COVID-19 according to WHO interim guidance | Critical cases | 55 | 47.7 (SD 15.49) | 58.2 | 24.62 (SD 3.31) | 15 (SD 6.32) | 25.45 | NR |
| Garrigues et al | 2020 | France | cohort | 110.9 | hospitalized in COVID-19 ward unit | Deceased, unreachable, demented, bedridden and non-French speaking patients.Patients who were directly admitted to the ICU without being hospitalized in COVID-19 unit | 120 | 63.2(SD 15.7) | 62.5 | NR | 11.2 (SD 13.4) | NR | 20 |
| Boari et al | 2021 | Italy | cohort | 112 | (1) confirmed COVID-19 infection as determined by a positive RT-PCR swab; (2) bilateral pulmonary interstitial opacities on chest imaging; (3) ARDS | died | 94 | NR | NR | NR | NR | NR | NR |
| Carenzo L.;Protti A et al | 2021 | Italy | cohort | 168 | NR | NR | 47 | 59 (SD 10) | 79.0 | 28.4 (5.3) | 29 (IQR 19-35) | NR | 100 |
| Carenzo,L.;Dalla Corte,F. et al | 2021 | Italy | cohort | 168 | NR | NR | 71 | 57 (IQR 51–62) | 78.9 | 27.8 (IQR 24.9–34.5) | 31 (IQR 19–56) | NR | NR |
| Damanti et al | 2021 | Italy | cohort | 168 | treated with CPAP without previous intubation. | Patients chronically receiving CPAP for obstructive sleep apnoea; requiring ICU during the same admission; or enrolled in a concomitant randomized trial on the use of early CPAP; or with severe contraindications to CPAP | 67 | 62.8 (SD 10.77) | 85.0 | 28.5 (IQR 25.1 – 32.0) | 20.4 (SD 13.28) | NR | NR |
| DE Lorenzo et al | 2021 | Italy | cohort | 89 | adult patients with confirmed sars cov 2 | NR | 251 | 61.8 (IQR 53.5-70.7) | 71.3 | 27.8 (IQR 25-31.3) | 12 (IQR 7-23) | NR | 10.8 |
| Froidure et al | 2021 | Belgium | cohort | 95 | [severe and critical COVID-19 patients admitted and underwent a three-month follow-up in our hospital. all patients for whom at least two out of three (clinical, PFT, HRCT) follow-up evaluations were available](https://www.covid19treatmentguidelines.nih.gov/overview/) | the delay between admission and follow-up evaluation was lower than 60 days or exceeded 120 days. | 134 | 60 (IQR 53–68) | 59.0 | NR | 11 (Range 1-183) | NR | 22 |
| Frontera,J. et al | 2021 | USA | cohort | 168 | age ≥ 18 years, hospital admission, RT-PCR positive SARS-CoV2 infection, survival to discharge. | negative or missing SARS-CoV-2 RT-PCR test, or evaluation in an outpatient or emergency department setting only. Readmissions were excluded | 196 | 68 (IQR 55–77) | 65.0 | 26 (IQR 23− 30) | 10.3 (IQR 4.8–32.5) | NR | 35 |
|  |  |  |  |  |  |  | 186 | 69 (IQR 57–78) | 65.0 | 28 (IQR 24–35) | 8.3 (IQR 3.7–19.9) | NR | 29 |
| Gamberini et al | 2021 | Italy | cohort | 90 | (a) age>18 years; (b) admission to ICU due to respiratory failure determined by SARS-CoV-2 infection confrmed by RT-PCR swab; (c) need of invasive mechanical ventilation during ICU stay. | SARS-CoV2 positive patients admitted to the hospital for other reasons (i.e., trauma, stroke) or refusal to participate to the study. | 205 | 63 (IQR 55–70) | 72.7 | 28 (IQR 26–31) | 42 (IQR 31–57) | NR | 100 |
| Gautam et al. | 2021 | UK | cohort | 143.4*^#^ | hospital admission > 3 days, with fraction of inspired oxygen > 40% for >6 hours, new stroke, pulmonary embolism, deep venous thrombosis, delirium, elevated troponin levels, residual acute kidney injury, or tachycardia > 100bpm at discharge. | Patients with mild to moderate disease who did not meet any of the above disease severity criteria, frailty score 6 or above on admission or discharged to residential/nursing home | 200 | 56.5 (SD 13.2) | 62.5 | NR | 22.7 (SD 18.4) | 100 | 43.5 |
| Ghosn et al | 2021 | France | cohort | 177 | Hospitalized patients with a virologically confirmed COVID-19 | NR | 1137 | 61 (IQR 51- 71) | 63.0 | NR | 9 (IQR 5- 15) | 72 | 29 |
| Gianella et al | 2021 | Switzerland | cohort | 84 | 39 consecutive laboratory-confrmed COVID19 patients with pathological fndings on a chest CT scan | age<18 years, pregnancy and absence of a written informed consent. | 39 | 62.5 (IQR 51.3–71) | 76.9 | NR | 15 (IQR 12–22) | NR | 25.6 |
| Guler et al | 2021 | Switzerland | cohort | 128^#^ | adults who survived either mild to moderate or severe to critical COVID-19. | NR | 66 | 60.3 (SD 12.0) | 60.6 | 29.8 (SD 5.7) | NR | NR | NR |
|  |  |  |  |  |  |  | 47 | 52.9 (SD 10.9) | 57.4 | 25.5 (SD 4.7) |  |  |  |
| Parker et al | 2021 | UK | cohort | 76.3* | NR | NR | 36 | 52.5 (SD 11.4) | 64.0 | NR | 40 (IQR 29, 53) | NR | 100 |
| Shang et al | 2021 | China | cohort | 168 | Confirmed COVID-19 positive by RT-PCR tests for SARS-CoV-2. All patients were discharged and followed up by telephone for 6 months | NR | 796 | 62.0 (IQR 51.0-69.0) | 50.8 | NR | 21.0 (IQR 14.0-27.0) | NR | 4.8 |
| Spinicci et al | 2021 | Italy | cohort | 60 | NR | patients discharged for more than 10 weeks, patients unable to attend the visit because of hospitalization or residents in care facilities. | 100 | 67.5 (IQR 56–78.5) | 59.0 | NR | 16 (IQR 8–27) | 90% | 31 |
| Todt et al | 2021 | Brazil | cohort | 84 | ≥18 years-old admitted with microbiological confirmation of SARS-CoV-2 infection by RT-PCR who survived to hospital discharge. | NR | 251 | 53.6 (SD± 14.9) | 59.8 | NR | 5 (IQR 3–10) | 76.1 | 16.3 |
| Vaes et al | 2021 | Netherlands | cohort | 77.7*^#^, 163.8*# | PCR confirmed | NR | 239 (62 hospitalised) | 53 (IQR 47.8-60) | 37.0 | 28.2 (24.8-32.6) | NR | NR | NR |
| Wu,Qian.;Hou,X et al | 2021 | China | cohort | 168 | NR | patients who (1) died before the follow-up, (2) refused to participate in the follow-up, and (3) left the local area and could not complete the follow-up. | 132 | 42 (IQR 32-54) | 55.0 | NR | NR | NR | NR |
| Wu,Xiaojun et al | 2021 | China | cohort | 98, 189, 275, 348 | At least 18 years with severe COVID-19 discharged Hospital | Patients with a history of hypertension, diabetes, cardiovascular disease, cancer, and chronic lung disease,, or a history of smoking. Patients who required intubation and mechanical ventilation. | 83 | 60 (IQR 52-66) | 57.0 | 25.0 (IQR 23.5–27·.) | 29 (IQR 25-35) | 100 | NR |
| Evans et al | 2021 | UK | cohort | 140 | patients >18 yrs old discharged following admission to a medical assessment or ward for confirmed or clinician-diagnosed COVID-19. | had a confirmed diagnosis of a pathogen unrelated to the objectives of this study, or were not admitted, or had another life-limiting illness with life expectancy less than six months. | 1077 | 57.9 (SD 13) | 64.3 | 30.1 (IQR 26.8 - 34.5) | 9 (IQR 4 - 21) | 79 | NR |

**Supplementary Table 3**. Study patient-reported outcome measures (PROMs). *Key: * Mean follow up time. Follow up time documented from symptom onset (#) or since diagnosis ($). mMRC=Modified medical research scale, 6MWT= 6-minute walk test, SPPB= short physical performance battery, PCFS=post Covid-19 functional status, NYHA= New York Heart Association Classification, SGRQ= St George’s respiratory questionnaire, GOSe=Glasgow outcome scale, MFIQ= Multidimensional fatigue inventory, USCD= University of California, San Diego Dyspnoea scale, CFS= clinical frailty score, MMT=Manual muscle test, ISWT= Incremental shuttle walk test. EQ-5D= European Quality of Life Five Dimension, SF-36=Short Form 36 Health Survey Questionnaire, MRS= Modified Rankin score, SF-12=Short Form 12 Health Survey Questionnaire, HRQOL= Health related Quality of life.*

|  |  |  | |  | | Outcomes | | | | | | | | |
| --- | --- | --- | --- | --- | --- | --- | --- | --- | --- | --- | --- | --- | --- | --- |
|  |  | |  | |  | | Respiratory/General functioning PROMs | | | Quality of life | | | Return to work | Healthcare utilisation |
| Study | Median follow up from discharge (days) | | ITU admission only | | Symptom | | mMRC | 6MWT | other | EQ-5D | SF-36 | Other |  |  |
| Bellan et al | 98 | |  | | x | |  |  | SPPB |  |  |  |  |  |
| Du et al | 168 | |  | |  | |  |  | PCFS |  |  |  |  |  |
| Fayol et al | 168 | |  | | x | |  |  | NYHA |  |  |  |  |  |
| Gonzalez et al | 84 | | yes | | x | | x | x |  |  |  | SF-12 |  |  |
| Huang et al | 153 | |  | | x | | x | x |  | x |  |  |  |  |
| Latronico et al 3 months | 84 | | yes | |  | |  | x | Barthel Index |  | x |  | x |  |
| Latronico et al 6 months | 168 | | yes | |  | |  | x | Barthel Index |  | x |  |  |  |
| Lerum et al | 83 | |  | |  | | x | x |  | x |  |  |  |  |
| Leth et al | 84 | |  | | x | | x |  |  |  |  |  |  |  |
| Meng et al | 84,164 | |  | | x | | x |  |  |  |  | SGRQ |  |  |
| Menges et al | 196^$^ | |  | | x | | x |  |  | x |  |  | x | x |
| Miyazato et al | 108* | |  | | x | |  |  |  |  |  |  |  |  |
| Monti et al | 61 | | yes | |  | |  |  | Borg scale, GOSe | x |  |  | x |  |
| Morin et al | 113 | |  | | x | | x | x | Nijmegen Score, MFIQ |  | x |  |  |  |
| Munblit et al | 217.5 | |  | | x | | x |  |  | x |  |  |  |  |
| Qin et al | 84 | |  | | x | |  |  |  |  |  |  |  |  |
| Shah et al | 81.9*^#^ | |  | | x | |  | x | UCSD dyspnoea scale |  |  |  |  |  |
| Sigfrid et al | 222 | |  | | x | | x |  |  | x |  | Washington group domain |  |  |
| Smet et al | 74* | |  | | x | |  |  |  |  |  |  |  |  |
| SuarezRobles et al | 90 | |  | | x | |  |  |  |  |  |  |  | x |
| Sykes et al | 113 | |  | | x | | x |  |  | x |  |  |  |  |
| Taboada et al | 168 | | yes | | x | |  |  | PCFS | x |  |  |  |  |
| van den Borst et al | 71* | |  | |  | | x | x | CFS |  | x |  |  |  |
| Walle-Hansen et al | 186 | |  | |  | |  |  | SPPB | x |  |  |  |  |
| Wong et al | 91^#^ | |  | | x | |  |  |  | x |  |  |  |  |
| Wu Qian et al | 168 | |  | | x | |  |  |  |  |  |  |  |  |
| Xiong et al | 168 | |  | | x | |  |  |  |  |  |  |  |  |
| Zhao et al | 97 | |  | | x | |  |  |  |  |  |  | x |  |
| Garrigues et al | 110.9 | |  | | x | | x |  |  | x |  |  | x |  |
| Boari et al | 112 | |  | | x | |  |  |  |  |  |  |  |  |
| Carenzo L.;Protti A et al | 168 | | yes | |  | |  |  |  | x |  |  |  |  |
| Carenzo,L.;Dalla Corte,F. et al | 168 | |  | |  | |  |  |  |  |  |  | x |  |
| Damanti et al | 168 | |  | |  | | x | x | Borg scale |  |  |  |  |  |
| DE Lorenzo et al | 89 | |  | |  | | x |  |  | x |  |  |  |  |
| Froidure et al | 95 | |  | | x | | x |  |  |  |  |  |  |  |
| Frontera,J. et al | 168 | |  | |  | |  |  | barthel index |  |  | MRS | x | x |
| Gamberini et al | 90 | | yes | |  | |  |  |  |  |  | HRQOL |  |  |
| Gautam et al. | 143.4*^#^ | |  | | x | |  |  | CFS | x |  |  |  |  |
| Ghosn et al | 177 | |  | | x | |  |  |  |  |  |  | x | x |
| Gianella et al | 84 | |  | |  | | x | x |  |  |  | SF-12 |  |  |
| Guler et al | 128^#^ | |  | |  | | x | x |  |  |  |  |  |  |
| Parker et al | 76.3* | | yes | |  | |  |  | Borg scale, STS, Grip strength |  | x |  |  |  |
| Shang et al | 168 | |  | | x | |  |  |  |  |  |  |  |  |
| Spinicci et al | 60 | |  | | x | |  |  |  |  |  |  |  | x |
| Todt et al | 84 | |  | |  | | x |  |  | x |  |  |  | x |
| Vaes et al | 77.7*^#^ | |  | |  | |  |  | PCFS | x |  |  | x |  |
| Wu,Qian.;Hou,X et al | 168 | |  | |  | | x |  | Borg scale, MMT |  |  |  |  |  |
| Wu,Xiaojun et al | 98,189,275,348 | |  | |  | | x | x |  |  |  |  |  |  |
| Evans et al | 140 | |  | | x | |  |  | Dyspnoea-12, SPPB, ISWT | x |  | Washington short set score | x |  |

**Supplementary Table 4**. Quality assessment using a combination of the Newcastle-Ottawa score and Downs and blacks checklist. {{16110 Downs,S.H. 1998; 16111 Peterson, J 2011

| Author | aim/objective of the study clearly described? | Patient characteristics described? | Site number | Representativeness of the cohort | Controls present? | loss to follow up documented? | main findings of the study clearly described? | Adjustments made for confounders? | Publication peer reviewed? |
| --- | --- | --- | --- | --- | --- | --- | --- | --- | --- |
|  | 1=yes, 0 =no | 1= Yes  0= no selection criteria | 1=multi-site  0=single site | 2=truly representative, an unselected sample.  1= selected cohort  0= No description | 1= Yes  0=no | 1=yes  0=no | 1=yes,  0=no | 1=yes  0=no | 1=yes  0= no |
| Bellan et al | 1 | 1 | 0 | 1 | 0 | 1 | 1 | 0 | 1 |
| Du et al | 1 | 0 | 0 | 0 | 0 | 1 | 0 | 0 | 0 |
| Fayol et al | 1 | 1 | 0 | 1 | 0 | 1 | 1 | 0 | 1 |
| Gonzalez et al | 1 | 1 | 1 | 2 | 0 | 1 | 1 | 0 | 1 |
| Huang et al | 1 | 1 | 0 | 1 | 0 | 1 | 1 | 0 | 1 |
| Latronico et al 3 months | 1 | 0 | 0 | 1 | 0 | 1 | 1 | 0 | 0 |
| Latronico et al 6 months | 1 | 0 | 0 | 1 | 0 | 1 | 1 | 0 | 0 |
| Lerum et al | 1 | 1 | 1 | 2 | 0 | 0 | 1 | 0 | 1 |
| Leth et al | 1 | 1 | 0 | 2 | 0 | 1 | 1 | 0 | 1 |
| Meng et al | 1 | 0 | 0 | 1 | 0 | 0 | 1 | 0 | 0 |
| Menges et al | 1 | 0 | 1 | 1 | 0 | 1 | 1 | 0 | 0 |
| Miyazato et al | 1 | 0 | 0 | 2 | 0 | 1 | 1 | 0 | 1 |
| Monti et al | 1 | 0 | 0 | 1 | 0 | 1 | 1 | 0 | 1 |
| Morin et al | 1 | 1 | 0 | 1 | 0 | 1 | 1 | 0 | 1 |
| Munblit et al | 1 | 1 | 1 | 1 | 0 | 1 | 1 | 1 | 0 |
| Qin et al | 1 | 0 | 0 | 2 | 0 | 1 | 1 | 1 | 1 |
| Shah et al | 1 | 0 | 0 | 2 | 0 | 1 | 1 | 1 | 1 |
| Sigfrid et al | 1 | 1 | 1 | 2 | 0 | 1 | 1 | 1 | 0 |
| Smet et al | 0 | 0 | 0 | 0 | 0 | 0 | 1 | 0 | 1 |
| SuarezRobles et al | 1 | 0 | 0 | 0 | 0 | 0 | 1 | 0 | 1 |
| Sykes et al | 1 | 0 | 0 | 1 | 0 | 1 | 1 | 1 | 1 |
| Taboada et al | 1 | 0 | 1 | 1 | 0 | 1 | 1 | 0 | 1 |
| van den Borst et al | 1 | 1 | 0 | 1 | 0 | 1 | 1 | 1 | 1 |
| Walle-Hansen et al | 1 | 1 | 1 | 1 | 0 | 1 | 1 | 0 | 1 |
| Wong et al | 1 | 1 | 1 | 2 | 0 | 1 | 1 | 1 | 1 |
| Wu Qian et al | 1 | 1 | 0 | 1 | 0 | 1 | 1 | 0 | 1 |
| Xiong et al | 1 | 1 | 0 | 1 | 1 | 1 | 1 | 0 | 1 |
| Zhao et al | 1 | 1 | 1 | 1 | 1 | 1 | 1 | 0 | 1 |
| Garrigues et al | 1 | 1 | 0 | 1 | 0 | 1 | 1 | 1 | 1 |
| Boari et al | 1 | 1 | 0 | 1 | 0 | 0 | 1 | 0 | 1 |
| Carenzo L.;Protti A et al | 1 | 0 | 0 | 1 | 0 | 1 | 1 | 0 | 1 |
| Carenzo,L.;Dalla Corte,F. et al | 1 | 0 | 0 | 1 | 0 | 1 | 1 | 0 | 1 |
| Damanti et al | 1 | 1 | 0 | 1 | 0 | 1 | 1 | 0 | 1 |
| DE Lorenzo et al | 1 | 0 | 0 | 0 | 0 | 0 | 1 | 0 | 1 |
| Froidure et al | 1 | 1 | 0 | 1 | 0 | 0 | 1 | 0 | 1 |
| Frontera,J. et al | 1 | 1 | 1 | 1 | 1 | 1 | 1 | 0 | 1 |
| Gamberini et al | 1 | 1 | 1 | 1 | 1 | 1 | 1 | 0 | 1 |
| Gautam et al. | 1 | 1 | 1 | 0 | 0 | 0 | 1 | 0 | 1 |
| Ghosn et al | 1 | 0 | 1 | 1 | 0 | 1 | 1 | 0 | 1 |
| Gianella et al | 1 | 1 | 0 | 1 | 0 | 0 | 1 | 0 | 1 |
| Guler et al | 1 | 0 | 1 | 0 | 0 | 0 | 1 | 0 | 1 |
| Parker et al | 0 | 0 | 0 | 1 | 0 | 1 | 1 | 0 | 1 |
| Shang et al | 1 | 0 | 1 | 1 | 0 | 1 | 1 | 1 | 1 |
| Spinicci et al | 0 | 0 | 0 | 1 | 0 | 1 | 1 | 1 | 1 |
| Todt et al | 1 | 0 | 0 | 1 | 0 | 1 | 1 | 0 | 1 |
| Vaes et al | 1 | 0 | 1 | 1 | 0 | 1 | 1 | 0 | 1 |
| Wu,Qian.;Hou,X et al | 1 | 0 | 0 | 1 | 0 | 1 | 1 | 1 | 1 |
| Wu,Xiaojun et al | 1 | 1 | 0 | 0 | 0 | 1 | 1 | 1 | 1 |
| Evans et al | 1 | 1 | 1 | 0 | 0 | 1 | 1 | 1 | 0 |

**Supplementary Table 5.** Physical functional outcomes reported in individual studies.

| Author | Measure | Result |
| --- | --- | --- |
| Bellan et al | Short Physical performance battery (SPPB) | limited mobility (22.3%) |
| Evan et al | SPPB  Incremental shuttle walk test (ISWT) | 9.9 (SD 3.1)  Distance 436m (SD 260)  % predicted 46.2 (SD 358) |
| Parker et al | Sit to Stand in 60  Grip strength (kg) | 19.0 (8.4)  21.4 (9.7) |
| Walle-Hansen et al | SPPB | Group 1 10.6 (SD 2.5)  Group 2 7.8 (SD 3.1) |
| Wu,Qian.;Hou,X et al | Manual Muscle tests | Not normal 2% |

Note: Walle-Hansen et al Group 1 includes participants <75yrs old, Group 2 includes those >75yrs old.

**Supplementary Table 6**. Functional outcomes reported in the above studies.

| Author | Variable reported | Result |
| --- | --- | --- |
| Post covid functional status scale (PCFS). 0=complete functional outcome, 1= negligible limited function, 2=mild, 3=moderate, 4=severe | | |
| Du et al | % | Grade 0 = 71  Grade 1 = 9  Grade 2 = 13  Grade 3 = 7  Grade 4 = 0 |
| Taboada et al | % | Grade 0 = 31  Grade 1 = 24  Grade 2 = 22  Grade 3 = 18  Grade 4 = 6 |
| Vaes et al | Mean (SD) | 2.3 (1.1) |
| New York Heart Association (NYHA) | | |
| Fayol et al | % | Grade 2 or above: 56% |
| Barthel Index (out of 100, 100=independent) | | |
| Frontera et al | Median (IQR) | Population 1 95 (70 -100)  Population 2 100 (90-100) |
| Latronico et al | % | 3 months – 100  6 months- 100 |
| Clinical frailty scale (1-9) | | |
| Gautam et al | % | 1 5.2  2 20.6  3 34.1  4 29.0  5 6.7  6 3.7  7 0.7 |
| van den Borst et al | Divided into groups % | Not Frail- 85  Somewhat Frail- 4  Frail- 10 |
| Glasgow Outcome Scale extended (GoSE)- physical recovery and disability | | |
| Monti et al | % | Upper good recovery- 26  Lower good recovery- 41  Upper moderate disability- 18  Lower moderate disability- 13  Upper Severe- 3  Lower Severe- 0 |
| Multidimensional Fatigue Inventory questionnaire (MFIQ) (scale 1-5) | | |
| Morin et al | Median Score (IQR) | Reduced motivation 4.5 (IQR, 3.0-5.0)  Mental fatigue 3.7 (IQR, 3.0-4.5) |

Note: Frontera et al Population 1 represents their patient group with neurological symptoms. Population 2 includes their COVID

control group without neurological symptoms

**Supplementary Table 7:** Respiratory outcome measures reported in the included studies.

| Author | Variable Reported | Result |
| --- | --- | --- |
| Borg score (0-10, higher is more breathless) | | |
| Damanti et al | % | 0: 49%  1: 3%  2: 10%  3: 26%  4: 8%  5:3%  6: 3% |
| Monti et al | % | Nil 60%  Very light 6%  Light 8%  Moderate 2%  Intense 5%  Strong 2%  Very strong 2% |
| Parker et al | Mean (SD) | Mean 4.6 (SD 2.2) |
| Wu,Qian.;Hou,X et al | % | 0: 89%  ≥1: 11% |
| Nijmegen score (>22 is dysfunctional breathing) | | |
| Morin et al | % >22 | 21 % |
| UCSD dyspnoea score (scale 0-120) | | |
| Shah et al | Median (IQR) | 11 (IQR 3–26) |
| Dyspnoea-12 (0-36, higher =more breathless) | | |
| Evans et al | Mean (SD) | 6.3 (SD 8.6) |
| St. George's respiratory questionnaire (SGRQ). Higher score is more severe | | |
| Gianella et al | Mean (SD) | Total 17 (17.4) |
| Meng et al | Median (IQR) | Group A: 18 (15 -27)  Group B: 17 (16 – 20)  Group C: 9 (4 – 28) |

Note: Meng et al Group A represents 3 month follow up of moderate severity Covid, Group B represents 3-month follow-up of severe COVID, and Group C represents 6 month follow up of severe Covid.

**Quality of Life Outcome Measures**

**Supplementary table 8.** SF-36 (0-100).

| First Author | Variable reported | Subgroup | Physical functioning | Social functioning | Pain | Mental health | Physical role | Emotional role | vitality | General health |
| --- | --- | --- | --- | --- | --- | --- | --- | --- | --- | --- |
| Latronico et al 3 months | Median |  | 80 | 73 | 78 | 79 | 46 | 69 | 64 | 65 |
| Latronico et al 6 months | Median |  | 82 | 78 | 76 | 76 | 68 | 76 | 65 | 64 |
| Morin et al | Mean |  | 71.3 | 74.6 | 70 | 66.3 | 25 | 73.7 | 46.9 | 57.5 |
| van den Borst et al | Mean | Moderate  Severe  Critical | 64.3; 54.5; 69.8 | 72.5; 63.3; 69.8 |  | 84.3; 78.2; 77.2 | 23.7; 41.6; 40.3 | 76.5; 64.3; 67.7 | 63.7; 55.9; 56.2 | 61.3; 45.4; 51.8 |
| Parker et al | Mean |  | 54.5 | 62.2 | 50.8 | 73.3 | 31 | 43.7 | 46.9 | 51.2 |

Note: Van den borst et al is split into 3 groups representing Moderate, Severe and Critical Covid infection respectively.
