## Supplementary material for "Patient-reported respiratory outcome measures in the recovery of adults hospitalised with COVID-19: A systematic review and meta-analysis": Forest Plots

**Meta-analyses of symptoms by 4 months – Forest plots**

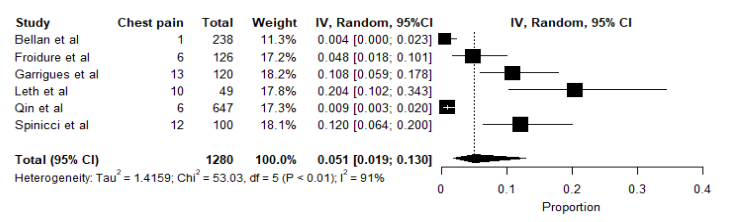

**Figure 1**. Meta-analysis of proportions of chest pain as a symptom up to 4 months after discharge. Black box, effect estimates from single studies; Diamond, pooled proportion with confidence interval; Weight (in %), influence an individual study had on the pooled result.

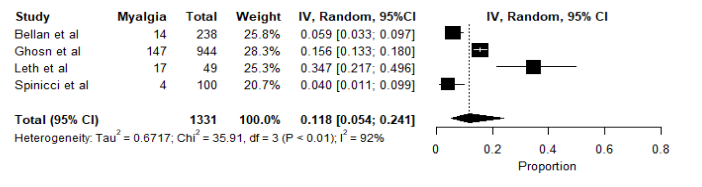

**Figure 2.** Meta-analysis of proportions of myalgia as a symptom up to 4 months after discharge. Black box, effect estimates from single studies; Diamond, pooled proportion with confidence interval; Weight (in %), influence an individual study had on the pooled result.

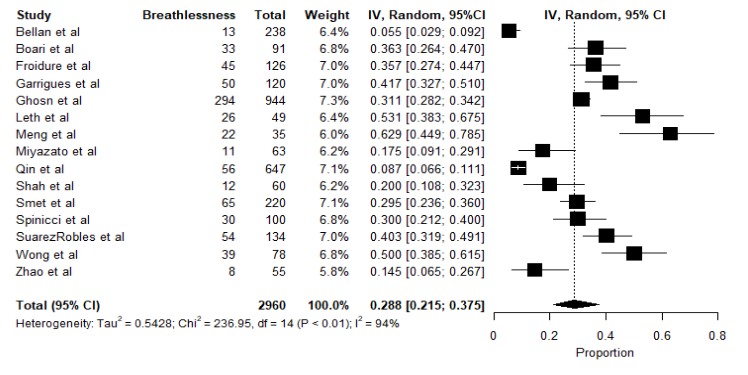

**Figure 3**. Meta-analysis of proportions of breathlessness as a symptom up to 4 months after discharge. Black box, effect estimates from single studies; Diamond, pooled proportion with confidence interval; Weight (in %), influence an individual study had on the pooled result.

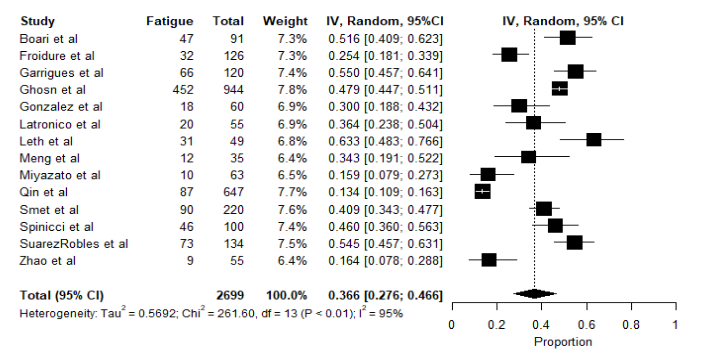

**Figure 4**. Meta-analysis of proportions of fatigue as a symptom up to 4 months after discharge. Black box, effect estimates from single studies; Diamond, pooled proportion with confidence interval; Weight (in %), influence an individual study had on the pooled result.

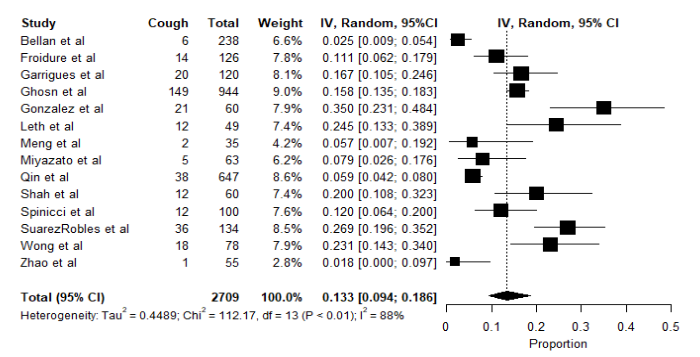

**Figure 5**. Meta-analysis of proportions of cough as a symptom up to 4 months after discharge. Black box, effect estimates from single studies; Diamond, pooled proportion with confidence interval; Weight (in %), influence an individual study had on the pooled result.

**Meta-analyses of symptoms over 4 months – Forest plots**

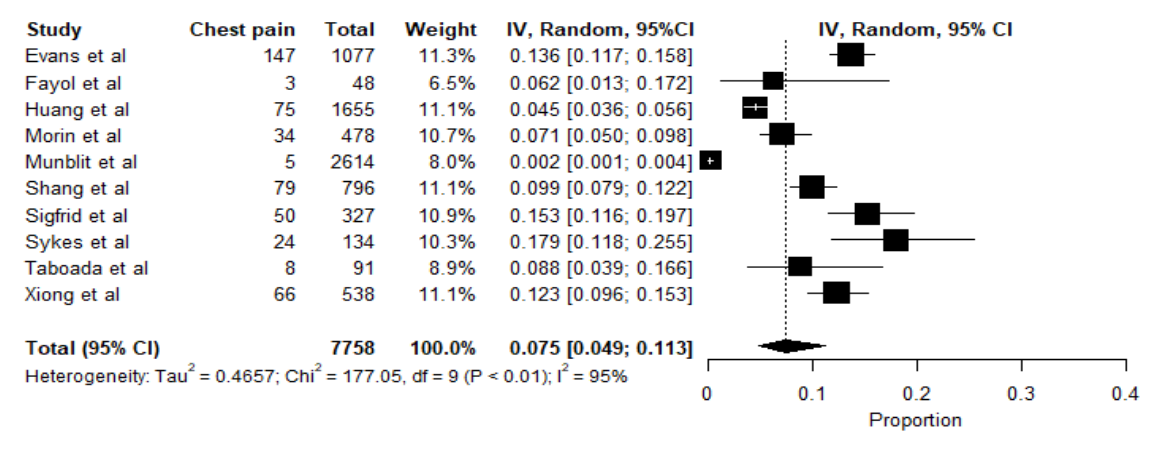

**Figure 6.** Meta-analysis of proportions of chest pain as a symptom over 4 months after discharge. Black box, effect estimates from single studies; Diamond, pooled proportion with confidence interval; Weight (in %), influence an individual study had on the pooled result.

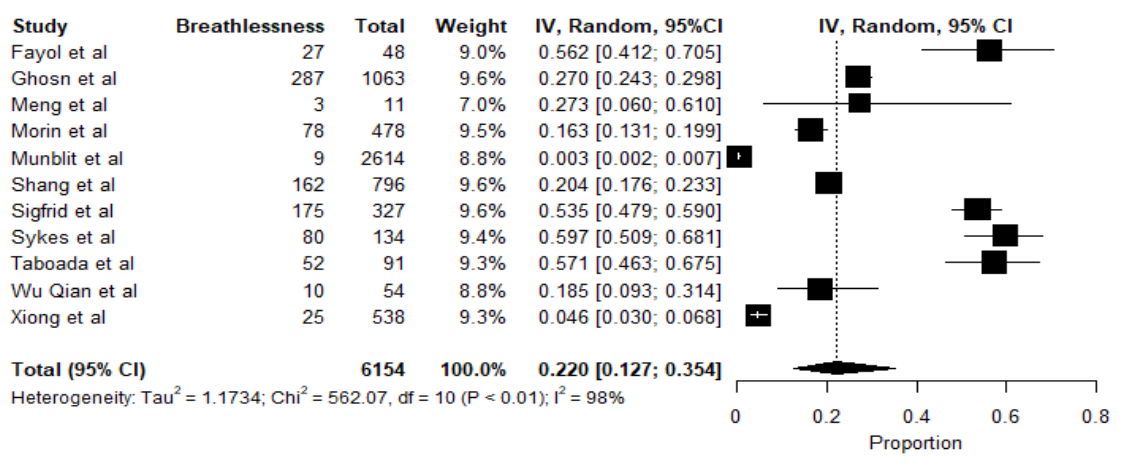

**Figure 7.** Meta-analysis of proportions of myalgia as a symptom over 4 months after discharge. Black box, effect estimates from single studies; Diamond, pooled proportion with confidence interval; Weight (in %), influence an individual study had on the pooled result.

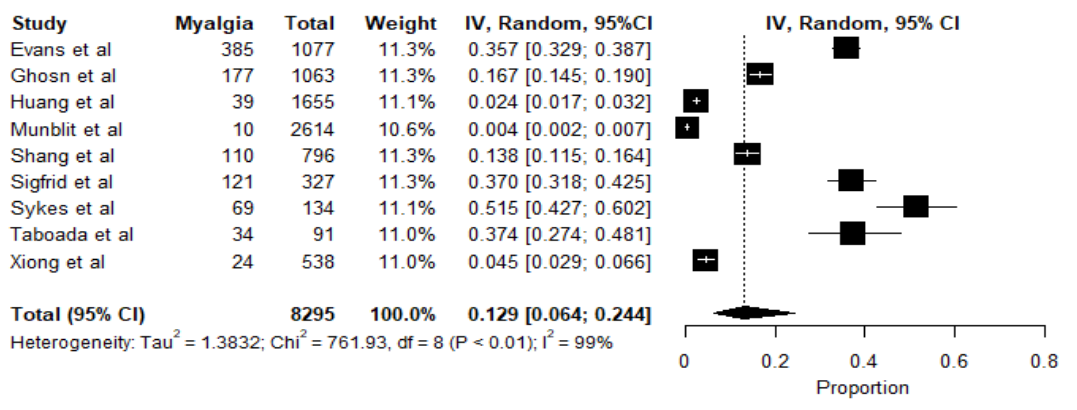

**Figure 8**. Meta-analysis of proportions of breathlessness as a symptom over 4 months after discharge. Black box, effect estimates from single studies; Diamond, pooled proportion with confidence interval; Weight (in %), influence an individual study had on the pooled result.

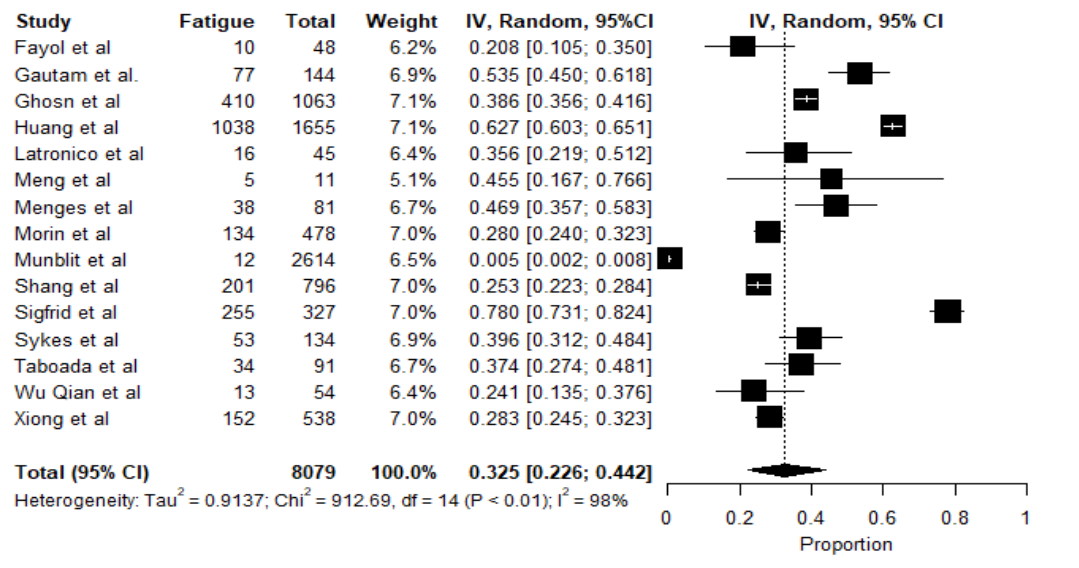

**Figure 9**. Meta-analysis of proportions of fatigue as a symptom over 4 months after discharge. Black box, effect estimates from single studies; Diamond, pooled proportion with confidence interval; Weight (in %), influence an individual study had on the pooled result.

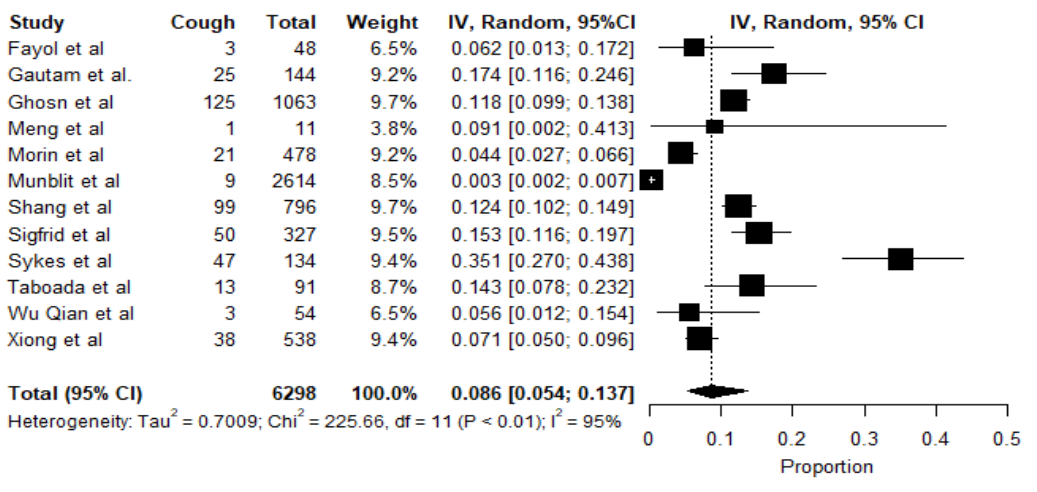

**Figure 10**. Meta-analysis of proportions of cough as a symptom over 4 months after discharge. Black box, effect estimates from single studies; Diamond, pooled proportion with confidence interval; Weight (in %), influence an individual study had on the pooled result.

**Meta-analyses of 6MWT by 4 months – Forest plot**

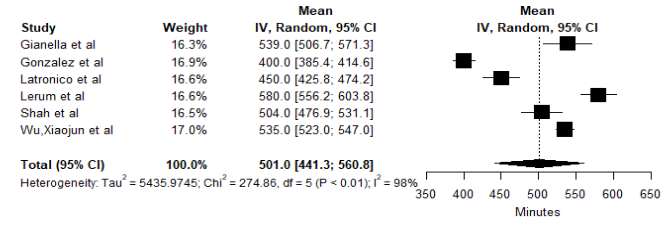

**Figure 11.** Meta-analysis of mean of 6MWT up to 4 months after discharge. Black box, effect estimates from single studies; Diamond, pooled proportion with confidence interval; Weight (in %), influence an individual study had on the pooled result.

**Meta-analyses of 6MWT over 4 months – Forest plot**

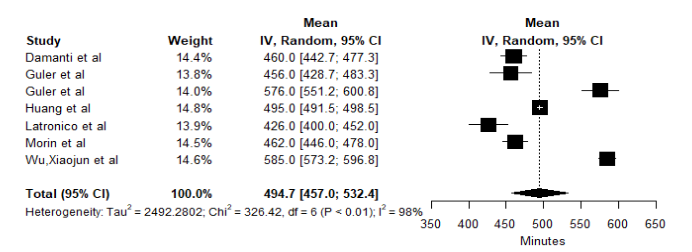

**Figure 12**. Meta-analysis of mean of 6MWT over 4 months after discharge. Black box, effect estimates from single studies; Diamond, pooled proportion with confidence interval; Weight (in %), influence an individual study had on the pooled result.

**Meta-analysis of SPPB over 4 months – Forest plot**

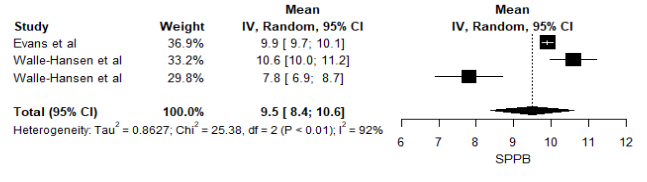

**Figure 13.** Meta-analysis of mean of SPPB over 4 months after discharge. Black box, effect estimates from single studies; Diamond, pooled proportion with confidence interval; Weight (in %), influence an individual study had on the pooled result.

**Meta-analyses of EQ-5D by 4 months– Forest plots**

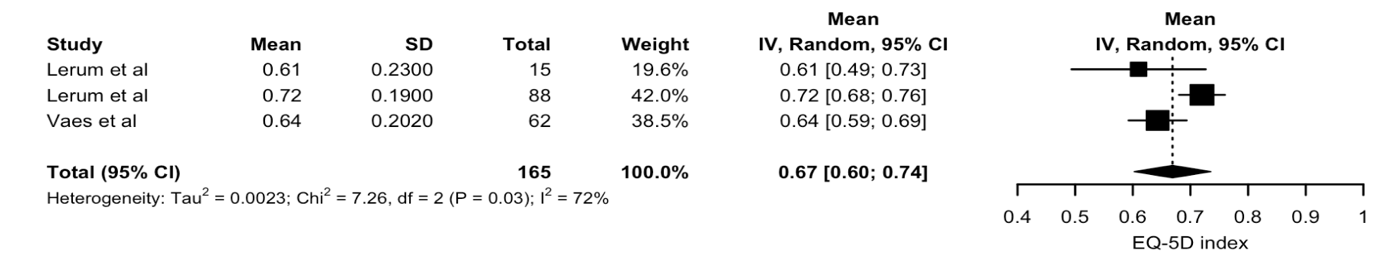

**Figure 14.** Meta-analysis of mean of EQ-5D index up to 4 months after discharge. Black box, effect estimates from single studies; Diamond, pooled proportion with confidence interval; Weight (in %), influence an individual study had on the pooled result.

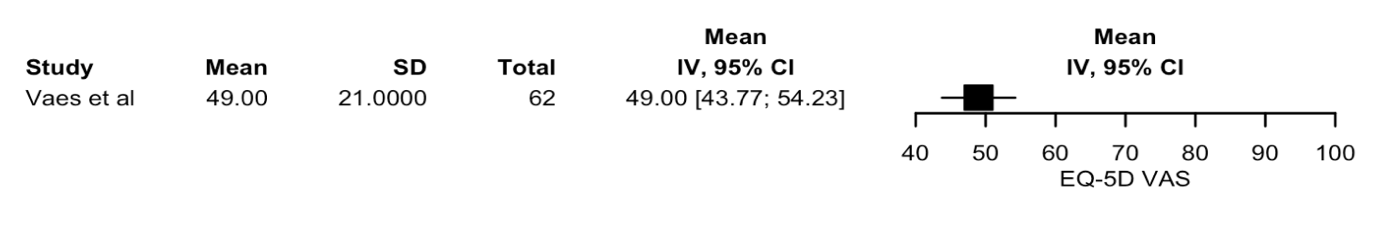
**Figure 15**. Meta-analysis of mean of EQ-5D VAS up to 4 months after discharge. Black box, effect estimates from single studies; Diamond, pooled proportion with confidence interval; Weight (in %), influence an individual study had on the pooled result.

**Meta-analyses of EQ-5D over 4 months– Forest plots**

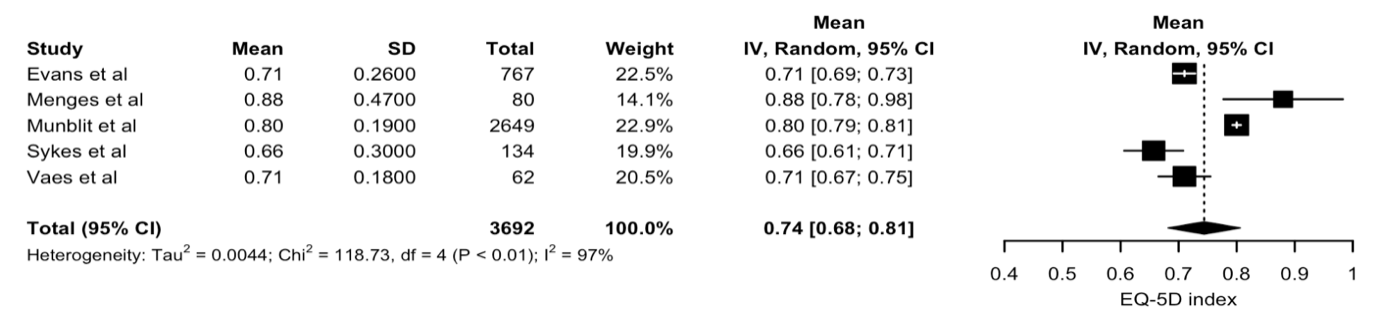

**Figure 16.** Meta-analysis of mean of EQ-5D index over 4 months after discharge. Black box, effect estimates from single studies; Diamond, pooled proportion with confidence interval; Weight (in %), influence an individual study had on the pooled result.

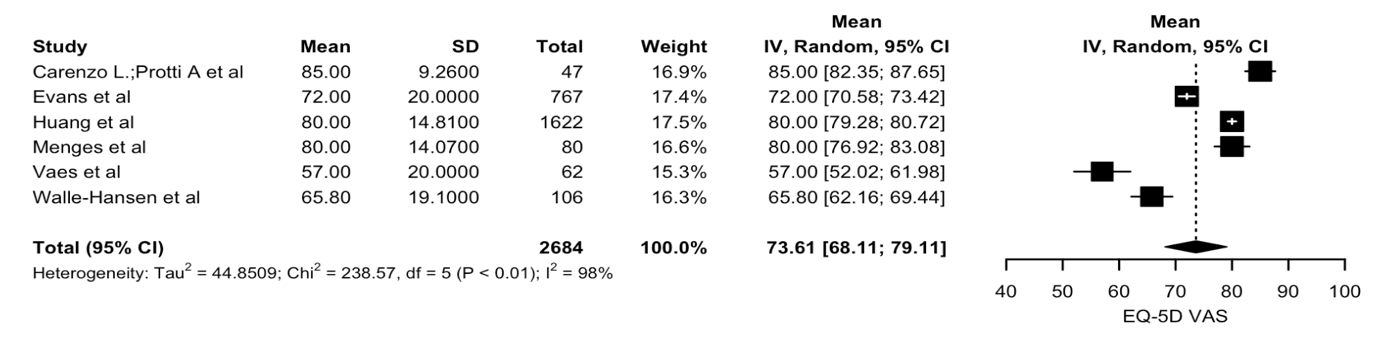
**Figure 17**. Meta-analysis of mean of EQ-5D VAS over 4 months after discharge. Black box, effect estimates from single studies; Diamond, pooled proportion with confidence interval; Weight (in %), influence an individual study had on the pooled result.

**Meta-analyses of mMRC score by 4 months– Forest plots**

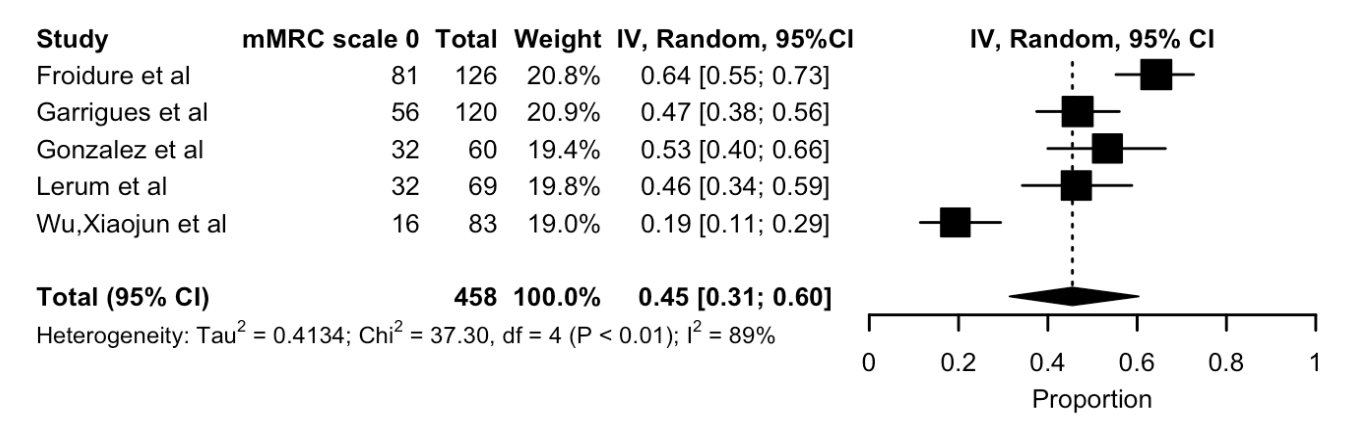

**Figure 18.** Meta-analysis of proportion of patients with a mMRC score equal to 0 up to 4 months after discharge. Black box, effect estimates from single studies; Diamond, pooled proportion with confidence interval; Weight (in %), influence an individual study had on the pooled result.

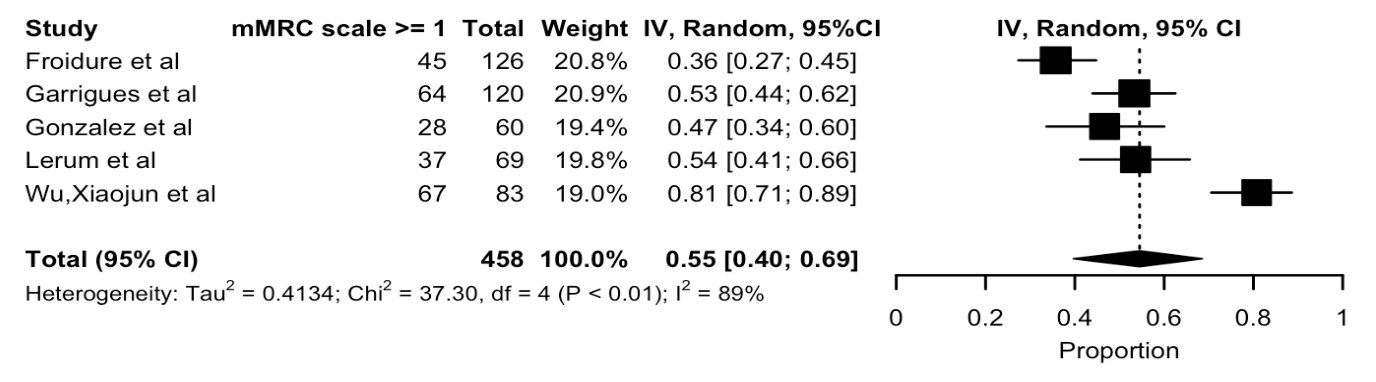

**Figure 19**. Meta-analysis of proportion of patients with a mMRC score greater than 0 up to 4 months after discharge. Black box, effect estimates from single studies; Diamond, pooled proportion with confidence interval; Weight (in %), influence an individual study had on the pooled result.

**Meta-analyses of mMRC score over 4 months– Forest plots**

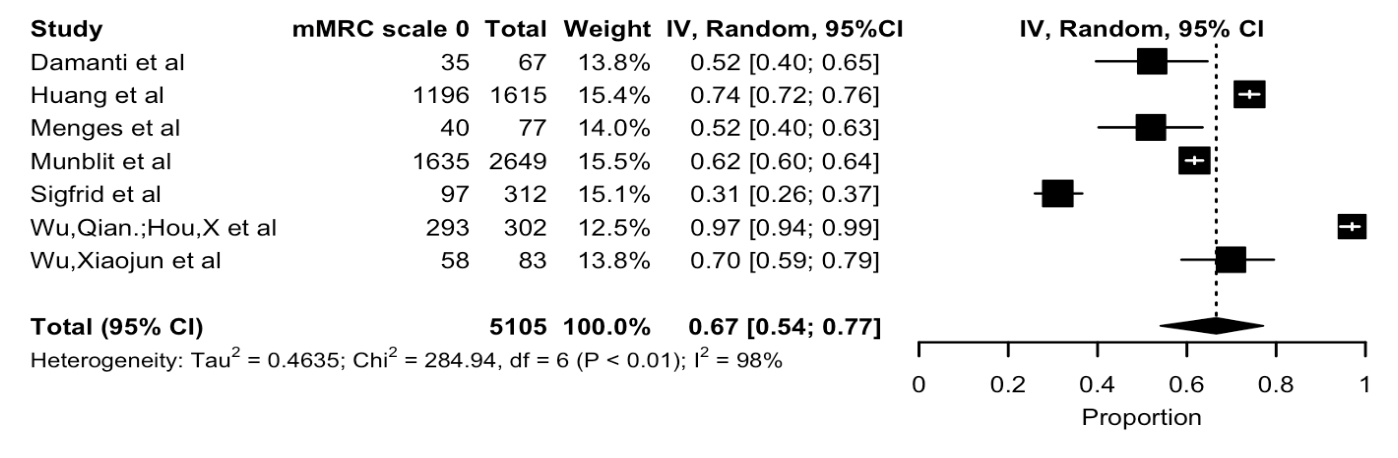

**Figure 20.** Meta-analysis of proportion of patients with a mMRC score equal to 0 over 4 months after discharge. Black box, effect estimates from single studies; Diamond, pooled proportion with confidence interval; Weight (in %), influence an individual study had on the pooled result.

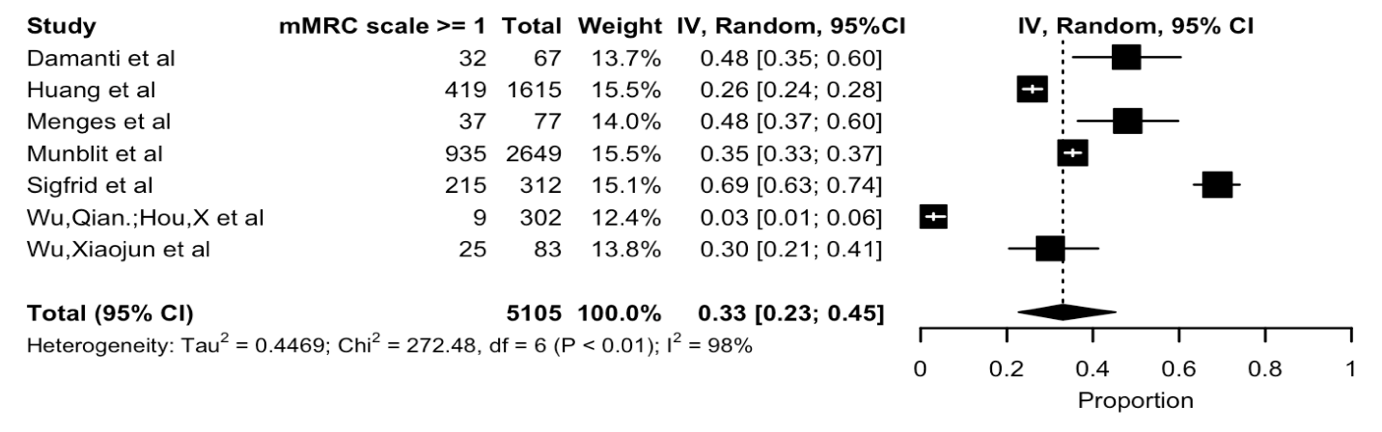

**Figure 21**. Meta-analysis of proportion of patients with a mMRC score greater than 0 over 4 months after discharge. Black box, effect estimates from single studies; Diamond, pooled proportion with confidence interval; Weight (in %), influence an individual study had on the pooled result.
